# Ornithine Decarboxylase (*ODC1*) gene variant (rs2302615) is associated with gastric cancer independently of *Helicobacter pylori* CagA serostatus

**DOI:** 10.1101/2021.04.13.21254467

**Authors:** Anna K Miller, Gloria Tavera, Ricardo L Dominguez, M Constanza Camargo, Tim Waterboer, Keith T Wilson, Scott M Williams, Douglas R Morgan

## Abstract

The primary cause of gastric cancer is chronic infection with *Helicobacter pylori (H. pylori)*, particularly the high-risk genotype *cagA*, and risk modification by human genetic variants. We studied 94 variants in 54 genes for association with gastric cancer, including rs2302615 in ornithine decarboxylase (*ODC1*), which may affect response to chemoprevention with the ODC inhibitor, eflornithine (difluoromethylornithine; DFMO). Our population-based, case-control study included 1366 individuals (664 gastric cancer cases and 702 controls) from Western Honduras, a high incidence region of Latin America. CagA seropositivity was strongly associated with cancer (OR = 3.6; 95% CI: 2.6, 5.1). The *ODC1* variant rs2302615 was associated with gastric cancer (OR = 1.36; p = 0.018) in a model adjusted for age, sex, and CagA serostatus. Two additional single nucleotide polymorphisms (SNPs) in *CASP1* (rs530537) and *TLR4* (rs1927914) genes were also associated with gastric cancer. The *ODC1* SNP association with gastric cancer was stronger in individuals who carried the TT genotype at the associating *TLR4* polymorphism, rs1927914 (OR = 1.77; p = 1.85 × 10^−3^). In conclusion, the *ODC1* variant, rs2302615, is associated with gastric cancer and supports chemoprevention trials with DFMO, particularly in individuals homozygous for the risk allele, C, at rs2302615.

## INTRODUCTION

Gastric adenocarcinoma is the third leading cause of global cancer mortality, and the leading infection-associated cancer, driven by chronic infection with *Helicobacter pylori*.(1, 2) There is also remarkable geographic variability of gastric cancer (GC) with respect to both incidence(3) and mortality.(4) Latin America has among the highest GC incidence rates in the world. There is an excess burden in the mountainous regions along the Pacific littoral as compared with coastal populations, even though the two regions have high endemic *H. pylori* infection; this has been termed the “Latin American altitude enigma”.(3) Altitude appears to be a surrogate for the clustering in the mountain villages of high-risk interactions between host genetic and *H. pylori* genetic factors(5), with modulation by dietary and environment factors.(6)

In the United States, GC represents a marked cancer disparity, with an excess burden among minorities. The GC incidence is approximately doubled among non-whites, including Hispanics.(7) Notably, immigrants from high GC incidence areas are at-risk. Those who emigrate from high-to-low incidence regions maintain the risk of their nation of origin, most likely due to their ‘importing’ their host and *H. pylori* genetic risks.(8)

The gastric carcinogenesis pathway is a multifactorial process that progresses from pre-malignant to malignant phenotypes through several histopathology stages: normal mucosa, chronic gastritis, atrophic gastritis, intestinal metaplasia, dysplasia, and adenocarcinoma.(9) Progression is driven by host genetics and *H. pylori* virulence and oncogenic factors, as well as dietary and environmental influences. *H. pylori* accounts for much of the attributable risk with *cagA* being the principal *H. pylori* risk genotype, and the CagA protein as the dominant virulence factor for gastric adenocarcinoma.(10-14)

Human germline mutations are thought to be important drivers in up to 10-15% of incident GC cases.(15) An elevated risk of GC has been associated with gene polymorphisms including those in the inflammation pathway, such as genes encoding IL-1β, TNF-α, and IL-10.(12, 16, 17) Additionally, associations between GC and variants in *PSCA* and *MUC1* (18-20), and at a locus that includes *PRKAA1* and *PTGER4* (20, 21) have been detected in genome-wide association studies of GC in Asian populations. Few studies have explicitly examined the host genetic basis for GC in Latin American populations.(22) The interaction of genetic variants in both the host and *H. pylori* affect disease progression and may explain some of the geographic variation in GC risk, yet the specific human loci and their relationship to disease risk remain poorly understood.(5, 23, 24)

Knowledge of human cancer risk loci may identify at-risk populations and has potential for targeting chemoprevention. For example, in colorectal cancer with the adenoma precursor lesion, there are potential targeted treatments for chemoprevention such as those based on ornithine decarboxylase (*ODC1*) gene variants. In humans, an *ODC1* variant, rs2302615, has been associated with adenoma risk, as well as an augmented chemoprevention response to alpha-difluoromethylornithine (DFMO). Patients with the CC genotype have a higher risk of colon adenomas, yet are more responsive to DFMO and sulindac.(25, 26) This *ODC1* SNP is located in intron 1, a region known to affect *ODC1* transcription (25), but no data exists on the relationship of this polymorphism and human GC. However, this gene may also be relevant to GC as myeloid-cell specific deletion of the *Odc1* gene in mice results in enhanced host immune response to *H. pylori* and diminished bacterial load in the stomach.(27) *H. pylori*-induced ODC activity is associated with macrophage apoptosis.(28, 29) In addition, studies have identified variants in ODC pathway genes that associate with GC.(30) Based on these prior studies, we hypothesized that *ODC1*, and rs2302615 in particular, which associates with transcription, will affect the risk for GC.

In addition to evaluating the *ODC1* variant described above, we analyzed 93 candidate single nucleotide polymorphisms (SNPs) in 53 genes potentially linked to GC based on prior association studies of gastric cancer and gastritis.(31-37) These markers have largely been studied in European and East Asian populations, with few studies in high risk Latin American populations. They fall into classes related to mechanisms of inflammation, immunity, *H. pylori* colonization, and oxidative stress (38-42), as well as many of the dietary and environmental markers associated with disease, but some do not have a clear mechanism.(6, 43)

Our study aimed at determining patterns of genetic association in the mountainous regions of Western Honduras, where the GC incidence is among the highest in Latin America.(3) We assessed if the genotypes of the *ODC1* SNP, rs2302615, were associated with GC in the high-risk Western Honduran population and in what context they were the most strongly associated. We also examined the other SNPs with known significance in GC and asked if putative association of the other SNPs was modified by the *ODC1* genotype risk.

## RESULTS

Of the 1500 individuals in this population-based, case-control study in Honduras, complete genetic data were available for 1366 participants. The mean ages of GC cases and controls were 63.8 and 53.6 years, respectively. Males comprised 70% (n= 468) and 50% (n= 350) of cases and controls, respectively. *H. pylori* and CagA seropositive cases were determined by a multiplex serology (see Methods). Overall, a high *H. pylori* prevalence was confirmed, detected in 90.5% and 88.2% of cases and controls, respectively. *H. pylori* CagA seropositivity was 87% and 73.6% in cases and controls, respectively, and strongly associated with GC in the unadjusted analysis (Odds Ratio, OR = 3.59; 95% CI 2.56, 5.11; p = 6.0 × 10^−14^; Table 1).

**Table 1.**
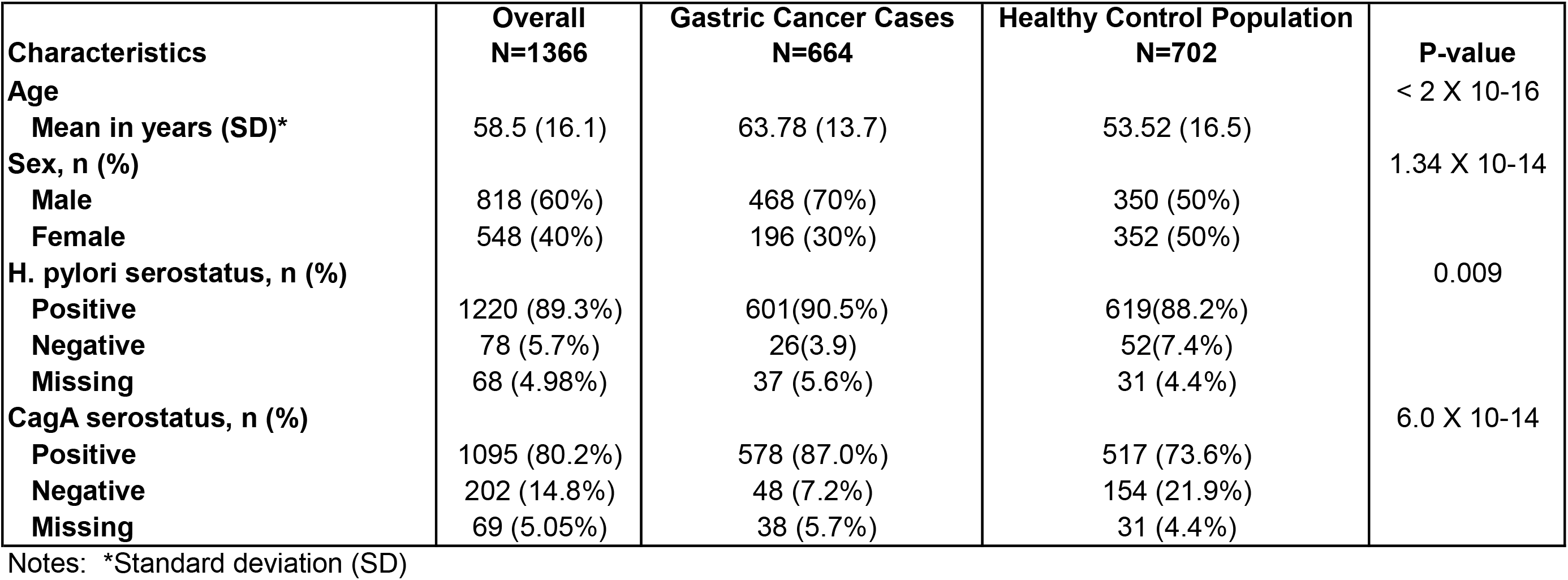
Selected characteristics of gastric cancer cases and population controls.

Of the 94 selected SNPs, one SNP, rs4072037, was out of Hardy–Weinberg equilibrium (p < 1.0 × 10^−16^ for both cases and controls), indicating a likely genotyping error, and was excluded from the analyses. In addition, 11 SNPs were excluded with a minor allele frequency of less than 0.05 (Table S1, Table S2). Linkage Disequilibrium (LD) was detected between 20 SNP pairs in which 4 SNPs appeared in two pairs. A total of 28 SNPs were excluded, resulting in 66 SNPs for the final analysis (Table S3, Figure S1). The Benjamini-Hochberg FDR test with an FDR level of 0.1 determined a multiple testing threshold of p = 0.021.

Cases were associated with *ODC1* genotype considered as cancer risk (CC) or protective (CT/TT) based on the colon cancer literature using a chi-square analysis (p = 0.021).(44-46) In the unadjusted logistic regression model, the *ODC1* SNP was associated with GC (OR=1.21, 95% CI 1.14, 1.80; p=0.027). In the model adjusted for age, sex, and CagA status, and FDR corrected for multiple testing, the *ODC1* SNP was significantly associated with GC (OR = 1.36, 95% CI 1.05, 1.76; p=0.018) (Table 2, Table S4).

**Table 2.** Associations of single nucleotide polymorphisms (SNPs) with gastric cancer risk.

| Chromosome | SNP | Gene | Major allele | Minor allele | MAF | Unadjusted OR (95% CI) | p-value (unadjusted) | Adjusted* OR (95% CI) | Adjusted p-value |
| --- | --- | --- | --- | --- | --- | --- | --- | --- | --- |
| 2 | rs2302615 | *ODC1* | C | T<sup>a</sup> | 0.264 | 1.21 (1.14, 1.80) | 0.027 | **1.36 (1.05, 1.76)** | **0.018** |
| 9 | rs1927914 | *TLR4* | T<sup>a</sup> | C | 0.365 | **1.33 (1.13, 1.54)** | **0.0004** | **1.23 (1.03, 1.47)** | **0.021** |
| 11 | rs530537 | *CASP1* | A<sup>a</sup> | G | 0.499 | **1.30 (1.11, 1.51)** | **0.0007** | **1.23 (1.03, 1.45)** | **0.019** |
| 2 | rs1056836 | *CYP1B1* | C<sup>a</sup> | G | 0.246 | **0.75 (0.63, 0.89)** | **0.001** | 0.81 (0.66, 0.99) | 0.036 |
| 2 | rs1800440 | *CYP1B1* | A<sup>a</sup> | G | 0.094 | **0.71 (0.55, 0.93)** | **0.011** | 0.74 (0.55, 0.98) | 0.04 |
| 3 | rs35683 | *GHRL* | C<sup>a</sup> | A | 0.276 | **0.81 (0.69, 0.96)** | **0.015** | 0.85 (0.71, 1.03) | 0.096 |
| 4 | rs4129009 | *TLR10* | A<sup>a</sup> | G | 0.107 | **0.73 (0.57, 0.94)** | **0.014** | 0.79 (0.59, 1.04) | 0.089 |
| 8 | rs2294008 | *PSCA* | T<sup>a</sup> | C | 0.322 | **0.81 (0.68, 0.95)** | **0.009** | 1.13 (0.74, 1.06) | 0.197 |

Notes:
- <sup>a</sup> referent allele - \* Adjusted for age, sex, and cagA serostatus, with respective ORs age (OR 1.03, 95% CI 1.02, 1.04), sex (OR 2.65, 95% CI 1.91, 3.71), and CagA (OR 3.16, 95% CI, 1.93, 5.26). - Odds ratio (OR) only included if p <0.10 and bolded SNPs pass FDR cut off

Of the remaining 65 SNPs, initial unadjusted association analyses identified seven SNPs with risk or protective associations with GC where the major allele was used as the reference. In an additive regression model, GC was associated with rs35686 in the ghrelin and obestatin prepropeptide (*GHRL*) gene, (p = 0.015), rs4129009 in toll-like receptor-10 (*TLR10*) (p = 0.014), prostate stem cell antigen (*PSCA*) (p = 0.009), rs1927914 in toll-like receptor-4 (*TLR4*) (p = 0.0004), and rs530537 in caspase-1 (*CASP1*) (p = 0.0007) (Table 2). Lastly, two SNPs, rs1056836 and rs1800440, in cytochrome P450 family 1 subfamily B member 1 (*CYP1B1*) were both associated with GC (p = 0.001 and p = 0.011, respectively) (Table 2). The two *CYP1B1* SNPs are not in LD (R^2^ = 0.034). In the models adjusted for age, sex, and bacterial CagA serostatus, only the SNPs in the *TLR4* and *CASP1* genes remained significant: *TLR4* (OR = 1.23; p = 0.021) and *CASP1* (OR = 1.23; p = 0.019).

In the final model with the three SNPs (*ODC1, TLR4, CASP1*) and adjusted for age, sex, and CagA serostatus, only the *ODC1* SNP remained significant (OR = 1.35; p = 0.021), as well as age (OR 1.03, 95% CI 1.02, 1.04), sex (OR 2.65, 95% CI 1.91, 3.71), and CagA (OR 3.16, 95% CI, 1.93, 5.26). SNPs in *TLR4* (OR = 1.04; p = 0.30) and *CASP1* (OR = 1.02; p = 0.27) were not significant in the final model. There were no detected interactions between *CASP1* and *ODC1* (OR = 1.02; p = 0.68) nor *TLR4* and *ODC1* (OR = 1.04; p = 0.52). However, noting the effects of the *TLR4* and *CASP1* loci on *ODC1*-related risk, we performed stratified analyses on the subsets of these two loci to assess how *ODC1* risk changes among strata. The effect of the *ODC1* genotype in the *TLR4* TT subset alone (n = 563; OR = 1.77; p =1.85 × 10^−3^) was significant, and with a larger effect size.

We also performed a stratified analysis restricting to the 1095 high-risk individuals who were CagA seropositive. Unadjusted analysis of these individuals were significant (p < 0.021) for the previously identified SNPs: rs1927914/*TLR4* (OR = 1.34; p = 0.005), rs1056836/*CYP1B1* (OR = 0.77; p = 0.008), and rs2302615/*ODC1* (OR = 1.35; p = 0.020). One additional SNP was identified in this high-risk cohort, rs4129009/*TLR10*, also passed the FDR threshold (OR = 0.717; p = 0.018). The ORs were of similar magnitude and size in the unadjusted and adjusted models for all individuals (Table 2, Table 3).

**Table 3.**
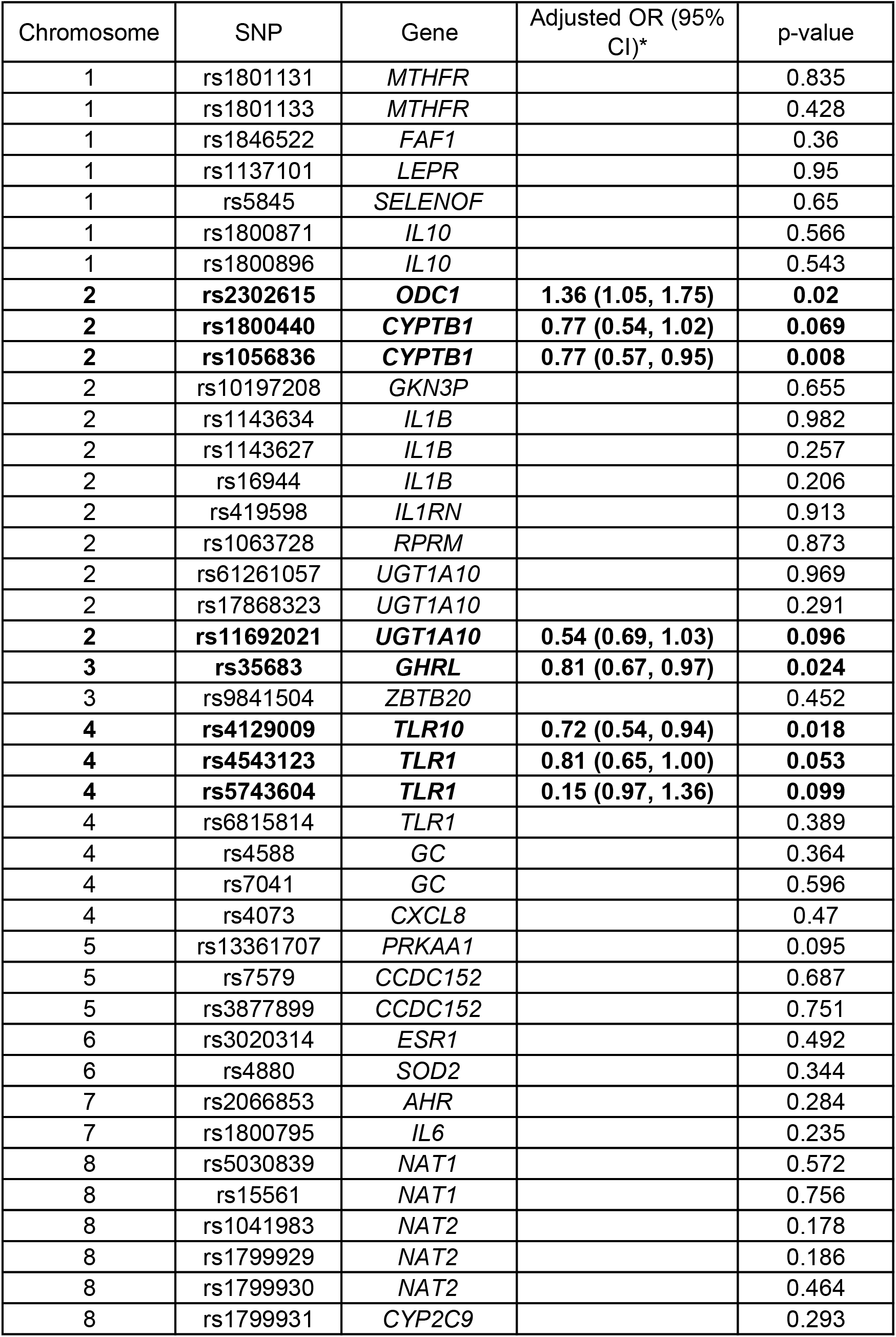
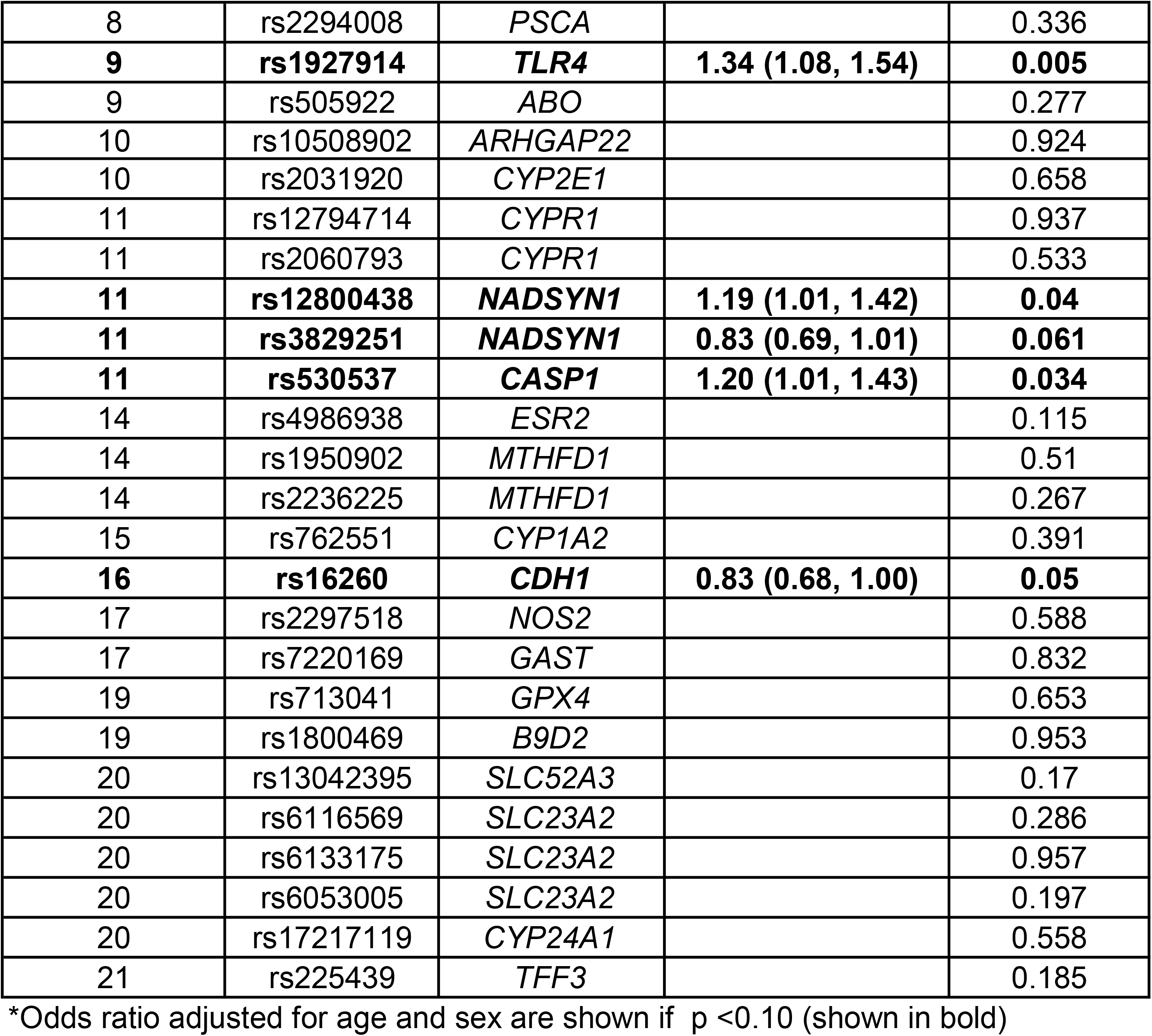
Associations of single nucleotide polymorphisms (SNPs) with gastric cancer risk among CagA-seropositive cases and controls.

## DISCUSSION

We found significant associations of the *ODC1* rs2302615 SNP, *TLR4* rs1927914, *CASP1* rs530537, and *H. pylori* CagA seropositivity with gastric adenocarcinoma in the rural high incidence region of Western Honduras. The *ODC1* rs2302615 SNP is of particular interest as it remained significant when adjusted for the other SNPs and is an actionable SNP previously seen in colorectal cancer.(25) CagA is the principal *H. pylori* oncogenic virulence factor for GC, and in studies with adjustment for CagA serostatus, the effect sizes of the human genetic variants are usually significantly reduced, (47, 48); however, in our study the effect sizes for *H. pylori* remained robust for the *ODC1* rs2302615 SNP following adjustment.

This is the first study, to our knowledge, demonstrating an association of an *ODC1* SNP with GC. Notably, in a Korean cohort investigation, other variants in this pathway were associated with GC, indicating that ODC may play a role in risk even if specific genes and variants differ by population.(21) In conjunction with the colon cancer and gut microbiota literature, these results indicate a broader role for this pathway in gastrointestinal cancers.(21, 49-54) ODC is the rate-limiting enzyme for polyamine biosynthesis that contributes to the pathogenesis of colitis and associated carcinogenesis by impairing M1 macrophage responses for antitumor immunity in a mouse model.(55) Loss of ODC in myeloid cells enhances host defense against *H. pylori*.(27) Similarly, EGFR signaling in macrophages has a significant role in colitis-associated carcinogenesis.(56) ODC generates polyamines that regulate the host immune response and is associated with DNA damage, due to the release of hydrogen peroxide by the back conversion of spermine to spermidine by spermine oxidase (SMOX), as spermidine and SMOX are associated with beta-catenin activation in gastric epithelial cells.(57-59) Both our current study and the colon cancer study by *Zell et al*. report odds ratios of similar magnitude for *ODC1* with the T allele as the reference allele: GC (OR = 1.36) and colorectal cancer survival (OR = 2.02).(45)

In addition to the *ODC1* SNP, rs2302615, we found that SNPs in the *CASP1* and *TLR4* genes were also associated with GC. The *ODC1* SNP genotype is highly significant when including covariates age, sex, CagA serostatus, and stronger in the *TLR4* TT subset. *CASP1* encodes for caspase-1, which is an important inflammasome mediator that cleaves precursors of key inflammatory cytokines into active proteins, including the interleukin-1 family and the pyroptosis inducer gasdermin D. (60-62) These *CASP-1*-dependent processes are critical in mediating an immune response that effectively dampens *H. pylori* infection and regulates the host immune response, especially to *cagA*-positive isolates.(63-68) *TLR4* encodes the Toll-like receptor-4, which elicits a strong inflammatory immune response by binding lipopolysaccharide ligands on the surface of Gram-negative bacteria and triggering the MyD88 inflammatory pathway to activate monocytes and neutrophils to clear infections.(69) Variants of *TLR4* have been associated with risk for chronic *H. pylori* infection in multiple studies in humans and mice.(70-72) They have also been implicated in carcinogenesis and metastasis and associated with a broad range of pathology, including recurrent spontaneous miscarriage and diabetic retinopathy in type-2 diabetes.(73, 74)

Our findings may have potential GC chemoprevention implications. The administration of DFMO to patients in high-risk GC regions where pre-malignant lesions are prevalent may be warranted. This concept is supported by findings that DFMO significantly attenuates GC development in *H. pylori*-infected Mongolian gerbils.(57, 58) If the mechanism is similar to colon cancer, DFMO could be used for chemoprevention in humans with a precision medicine approach and focused on those with the higher risk CC *ODC1* genotype, and especially in those who also have the *TLR4* TT genotype, which was observed in 40.6% of the patients in our population. The genotype CC *ODC1* and diplotype *ODC1* CC/ *TLR4* TT have a prevalence of 29.5% and 11.5%, respectively, in the Honduras population.

In summary, our findings suggest that *ODC1* genotyping and *H. pylori* CagA seropositivity is significantly associated with gastric adenocarcinoma in Western Honduras. In addition, its association and potential therapeutic efficacy may be enhanced in those with the TT genotype at *TLR4* rs1927914. Nonetheless, replication studies in diverse populations will be necessary to confirm possible generalizability. If confirmed by independent studies, this finding will be an important innovation in guiding a precision based intervention for GC in this and perhaps other high-risk populations and potentially reducing the GC mortality in the high incidence regions of Latin America and Asia.

## Subjects and Methods

### Study population

We conducted a population-based, case control study in the mountainous region of west-central Honduras. The population is of Hispanic-Mestizo ethnicity and this region has among the highest incidence rates in the western hemisphere, with a high prevalence of chronic *H. pylori* infection (80-90%).(75) Incident GC cases were enrolled prospectively from the two district hospitals (Santa Rosa de Copán, Siguatepeque) that serve the mountainous rural areas of west-central Honduras. The diagnosis of GC was based on endoscopic appearance and confirmatory histopathology. Household interviews were conducted for the randomly selected controls in the villages in the region. A novel multiplex serology determined *H. pylori* and CagA serostatus, the dominant bacterial risk genotype for GC.(37, 76, 77) We focused on CagA, determined by antibodies to this oncoprotein, as the optimal measure of cancer risk with bacterial chronic infection, given the high *H. pylori* prevalence in the region.

Of the 1500 people enrolled in the case-control study, complete genotyping results were available for 1366 subjects (664 cases, 702 controls). Overall, 89.3% and 80.2% of subjects were seropositive for *H. pylori* infection and CagA, respectively, as outlined in Table 1. SNPs were chosen in loci in distinct cancer risk pathways (Table S1), including the *ODC1* SNP (rs2302615) data that was analyzed both as one of the 94 SNPs and as a covariate, since it has postulated protective effects (CT/TT). The selected SNPs are biased towards coding regions with 17 non-coding SNPs in high LD with both coding and noncoding SNPs (R^2^ > 0.8) (Table S2, Table S3).

### Genetic variant analysis

Human DNA was isolated from whole blood samples with the Qiagen Puregene® kit and genotyped on the Sequenom® platform. The *ODC1* rs2302615 SNP was determined by TaqMan assay (Thermo Fisher). Some SNPs were also included as they previously associated with putative disease processes in colon cancer.(78) The 94 hypothesis-driven SNPs fell into four different categories, corresponding to functional SNPs in the GC literature: inflammation and immunity, oncogenic environmental factors, and nutrition (Table S1). (49-54)

### Statistical analyses

Using PLINK (version 1.9), 12 of the 94 selected SNPs were removed due to Hardy-Weinberg equilibrium or a minor allele frequency less than 0.05 (Table S1, Table S2).(79) Allele frequencies were within the expected ranges, based on the Latin American populations in the 1000 genomes databases.(80) False Discovery Rate (FDR) was used to calculate a threshold for individual SNP significance using the Benjamini-Hochberg test with a FDR level of 0.1 for all tests. In tests of significance across multiple models, trend tests were used including basic allelic test, the Cochran-Armitage trend test, dominant and recessive models and a genotypic test. Additionally, linkage disequilibrium (LD) was characterized in PLINK and Haploview (version 4.2) among the SNPs regardless of case-control designation to determine the number of independent tests for the FDR threshold.(81) A total of 66 SNPs were considered in the final analysis as linkage disequilibrium (LD) (R^2^ ≥ 0.9) was detected between 20 SNP pairs in which 4 appeared in two pairs, with 16 total SNPs excluded (Table S3, Figure S1).

Association tests and unadjusted analyses of the final 66 SNPs were performed in PLINK (version 1.9).(79) Analyses of the resulting SNPs significant at p < 0.1 were analyzed in adjusted models for covariates age, sex, CagA serostatus, and *ODC1* genotype were performed in R (version 3.6.3). In all cases except one, the referent allele was the major allele; the exception was *ODC1* where the prior work on colon cancer indicted that the CC genotype was the risk genotype, but in our population, C was the major allele. Therefore, we used the T allele as the referent for this SNP to assess the risk conferred by the CC genotype in parallel with the prior literature. Select covariates were included based on summary statistics in current data and prior studies. Covariates age and sex was expected to influence genetic effects as age positively associates with GC incidence.(82) GC incidence is also notably higher in males, revealing the possible impact of sex hormones on development of gastric disease.(83-85) CagA serostatus was included in all adjusted models as individuals exposed to cagA-positive bacteria associate with increased risk of GC severity.(47, 48) Stratified analyses were restricted to the 1095 individuals carrying *cagA*-positive bacteria to evaluate the effect of CagA using a univariate analysis. *ODC1* status was grouped into genotypes associated with putative risk (CC) or protection (CT/TT) based on previous studies.(25, 45) Logistic regression was used to estimate OR, adjusted for age, sex, CagA serostatus, and *ODC1* genotype. SNPs significant individually were also tested for interactions and assessed in stratified analyses.

## Data Availability

This data will be publicly available 6 months after formal publication.

## Acknowledgements

We recognize our colleagues in the Hospital de Occidente, Honduras Ministry of Health, and Central America Medical Outreach (CAMO), with special thanks to Lesby Castellanos, Carmen Ramos, Kathy Tschiegg, and Dr. Lia Suazo. We acknowledge the contributions of Dr. Enrique Martinez (Hospital Evangelico, Honduras) and Dr. Michael Pawlita (DKFZ).

