## Supplemental Table 1 for "Ornithine Decarboxylase (*ODC1*) gene variant (rs2302615) is associated with gastric cancer independently of *Helicobacter pylori* CagA serostatus"

**Table S1. Four categories of hypothesized gastric cancer etiologies for 94 study single nucleotide polymorphisms (SNPs)**

***1. Inflammation and Immunity***

| Gene | Chr | SNP |
|---|---|---|
| *MTHFR* | 1 | rs1801131 |
| *MTHFR* | 1 | rs1801133 |
| *IL10* | 1 | rs1800872 |
| *IL10* | 1 | rs1800871 |
| *IL10* | 1 | rs1800896 |
| *IL1B* | 2 | rs1143634 |
| *IL1B* | 2 | rs1143627 |
| *IL1B* | 2 | rs16944 |
| *TLR10* | 4 | rs4129009 |
| *TLR10* | 4 | rs10004195 |
| *TLR10* | 4 | rs12233670 |
| *TLR1* | 4 | rs4543123 |
| *TLR1* | 4 | rs4833095 |
| *TLR1* | 4 | rs5743604 |
| *TLR1* | 4 | rs17616434 |
| *TLR1* | 4 | rs6815814 |
| *CXCL8* | 4 | rs4073 |
| *IL6* | 7 | rs1800797 |
| *IL6* | 7 | rs1800795 |
| *NAT1* | 8 | rs15561 |
| *MBL2* | 10 | rs5030737 |
| *CYP2E* | 10 | rs2031920 |
| *MTHFD1* | 14 | rs1950902 |
| *MTHFD1* | 14 | rs2236225 |
| *B9D2* | 19 | rs1800469 |

***2. Oncogenesis***

| Gene | Chr | SNP |
|---|---|---|
| *FAF1* | 1 | rs1846522 |
| *LEPR* | 1 | rs1137101 |
| *SELENOF* | 1 | rs5845 |
| *MUC1* | 1 | rs4072037 |
| *GKN3P* | 2 | rs10197208 |
| *IL1RN* | 2 | rs419598 |
| *RPRM* | 2 | rs1063728 |
| *GHRL* | 3 | rs35683 |
| *ZBTB20* | 3 | rs9841504 |
| *PRKAA1* | 5 | rs13361707 |
| *CCDC152* | 5 | rs7579 |
| *CCDC152* | 5 | rs3877899 |
| *PGC* | 6 | rs1316979 |

| | | |
|---|---|---|
| *ESR1* | 6 | rs3020314 |
| *SOD2* | 6 | rs4880 |
| *SHBG* | 7 | rs799941 |
| *LOC105375494* | 7 | rs7799039 |
| *PSCA* | 8 | rs2294008 |
| *TLR4* | 9 | rs1927914 |
| *ABO* | 9 | rs8176747 |
| *ABO* | 9 | rs8176746 |
| *ABO* | 9 | rs505922 |
| *ARHGAP22* | 10 | rs10508902 |
| *CASP1* | 11 | rs530537 |
| *ESR2* | 14 | rs4986938 |
| *CDH1* | 16 | rs16260 |
| *NOS2* | 17 | rs2297518 |
| *GAST* | 17 | rs7220169 |
| *GPX4* | 19 | rs713041 |
| *TGFB1* | 19 | rs2241715 |
| *SLC52A3* | 20 | rs13042395 |
| *TFF3* | 21 | rs225439 |

***3. Environment and Nutrition***

| Gene | Chr | SNP |
|---|---|---|
| *EPHX1* | 1 | rs1801131 |
| *CYP1B1* | 1 | rs1051740 |
| *CYP1B1* | 2 | rs1800440 |
| *CYP1B1* | 2 | rs1056836 |
| *UGT1A10* | 2 | rs61261057 |
| *UGT1A10* | 2 | rs17868323 |
| *AHR* | 2 | rs11692021 |
| *NAT1* | 7 | rs2066853 |
| *NAT1* | 8 | rs5030839 |
| *NAT2* | 8 | rs4986782 |
| *NAT2* | 8 | rs1801279 |
| *NAT2* | 8 | rs1041983 |
| *NAT2* | 8 | rs1801280 |
| *NAT2* | 8 | rs1799929 |
| *NAT2* | 8 | rs1799930 |
| *NAT2* | 8 | rs1208 |
| *CYP2C9* | 8 | rs1799931 |
| *CYP1A2* | 10 | rs1057910 |

***4. Miscellaneous***

| Gene | Chr | SNP |
|---|---|---|
| *ODC1* | 2 | rs2302615 |
| *GC* | 4 | rs2282679 |
| *GC* | 4 | rs4588 |
| *GC* | 4 | rs7041 |

| | | |
|---|---|---|
| *GC* | 4 | rs1155563 |
| *LOC100419170* | 4 | rs11736691 |
| *SLC23A1* | 5 | rs33972313 |
| *CYP2R1* | 11 | rs1993116 |
| *CYP2R1* | 11 | rs10500804 |
| *CYP2R1* | 11 | rs12794714 |
| *CYP2R1* | 11 | rs2060793 |
| *NADSYN1* | 11 | rs7944926 |
| *NADSYN1* | 11 | rs12800438 |
| *NADSYN1* | 11 | rs3794060 |
| *NADSYN1* | 11 | rs3829251 |
| *SLC23A2* | 20 | rs6116569 |
| *SLC23A2* | 20 | rs6133175 |
| *SLC23A2* | 20 | rs6053005 |
| *CYP24A1* | 20 | rs17217119 |
