## Supplemental Table 2 for "Ornithine Decarboxylase (*ODC1*) gene variant (rs2302615) is associated with gastric cancer independently of *Helicobacter pylori* CagA serostatus"

**Table S2. Single nucleotide polymorphisms (SNPs) genotyped in this gastric cancer study**

| SNP | Chromosome | Gene | Location | Genic Location | MAF |
|---|---|---|---|---|---|
| rs1801131 | 1 | *MTHFR* | 11794419 | Missense | 0.137 |
| rs1801133 | 1 | *MTHFR* | 11796321 | Missense | 0.433 |
| rs1846522 | 1 | *FAF1* | 50515533 | Intron | 0.053 |
| rs1137101 | 1 | *LEPR* | 65592830 | Missense | 0.43 |
| rs5845 | 1 | *SELENOF* | 86863156 | Non Coding | 0.186 |
| rs4072037* | 1 | *MUC1* | 155192276 | Synonymous | 0.094 |
| rs1800872* | 1 | *IL10* | 206773062 | Upstream | 0.387 |
| rs1800871 | 1 | *IL10* | 206773289 | Upstream | 0.383 |
| rs1800896 | 1 | *IL10* | 206773552 | Upstream | 0.211 |
| rs1051740 | 1 | *EPHX1* | 225831932 | Missense | 0.386 |
| rs2302615 | 2 | *ODC1* | 10448012 | Intron | 0.264 |
| rs1800440 | 2 | *CYP1B1* | 38070996 | Missense | 0.094 |
| rs1056836 | 2 | *CYP1B1* | 38071060 | Missense | 0.246 |
| rs10197208 | 2 | *GKN3P* | 68925812 | none | 0.359 |
| rs1143634 | 2 | *IL1B* | 112832813 | Synonymous | 0.104 |
| rs1143627* | 2 | *IL1B* | 112836810 | 5' UTR | 0.388 |
| rs16944 | 2 | *IL1B* | 112837290 | Upstream | 0.391 |
| rs419598 | 2 | *IL1RN* | 113129630 | Synonymous | 0.349 |
| rs1063728 | 2 | *RPRM* | 153477985 | 3' UTR | 0.285 |
| rs61261057* | 2 | *UGT1A10* | 233682280 | Intron | 4E-04 |
| rs17868323 | 2 | *UGT1A10* | 233682324 | Intron | 0.44 |
| rs11692021 | 2 | *UGT1A10* | 233682559 | Intron | 0.23 |
| rs35683 | 3 | *GHRL* | 10286566 | Intron | 0.276 |
| rs9841504 | 3 | *ZBTB20* | 114643917 | Intron | 0.257 |
| rs4129009 | 4 | *TLR10* | 38773268 | Missense | 0.107 |
| rs10004195* | 4 | *TLR10* | 38783103 | Upstream | 0.162 |
| rs12233670* | 4 | *TLR10* | 38785595 | none | 0.195 |
| rs4543123 | 4 | *TLR1* | 38790903 | Downstream | 0.188 |
| rs4833095* | 4 | *TLR1* | 38798089 | Missense | 0.455 |
| rs5743604 | 4 | *TLR1* | 38799664 | Intron | 0.471 |
| rs17616434* | 4 | *TLR1* | 38811255 | none | 0.452 |
| rs6815814 | 4 | *TLR1* | 38814717 | none | 0.389 |
| rs2282679* | 4 | *GC* | 71742666 | Intron | 0.204 |
| rs4588 | 4 | *GC* | 71752606 | Missense | 0.206 |
| rs7041 | 4 | *GC* | 71752617 | Missense | 0.46 |
| rs1155563* | 4 | *GC* | 71777771 | Intron | 0.193 |
| rs4073 | 4 | *CXCL8* | 73740307 | Upstream | 0.376 |
| rs11736691* | 4 | *LOC100419170* | 153665441 | Intron | 0.014 |
| rs13361707 | 5 | *PRKAA1* | 40791782 | Intron | 0.208 |
| rs7579 | 5 | *CCDC152* | 42800706 | 3' UTR | 0.499 |
| rs3877899 | 5 | *CCDC152* | 42801166 | 3' UTR | 0.131 |
| rs33972313* | 5 | *SLC23A1* | 139379813 | Missense | 0.015 |
| rs1316979* | 6 | *PGC* | 41740926 | Intron | 0.031 |
| rs3020314 | 6 | *ESR1* | 151949537 | Intron | 0.395 |
| rs4880 | 6 | *SOD2* | 159692840 | Missense | 0.458 |

| | | | | | |
|---|---|---|---|---|---|
| rs2066853 | 7 | *AHR* | 17339486 | Missense | 0.153 |
| rs1800797* | 7 | *IL6* | 22726602 | Intron | 0.138 |
| rs1800795 | 7 | *IL6* | 22727026 | Intron | 0.139 |
| rs799941 | 7 | *SHBG* | 80431740 | none | 0.054 |
| rs7799039 | 7 | *LOC105375494* | 128238730 | Upstream | 0.406 |
| rs5030839* | 8 | *NAT1* | 18222606 | Stop Gained | 0.007 |
| rs4986782* | 8 | *NAT1* | 18222607 | Missense | 0.009 |
| rs15561 | 8 | *NAT1* | 18223142 | 3' UTR | 0.429 |
| rs1801279* | 8 | *NAT2* | 18400194 | Missense | 0.003 |
| rs1041983 | 8 | *NAT2* | 18400285 | Synonymous | 0.321 |
| rs1801280* | 8 | *NAT2* | 18400344 | Missense | 0.281 |
| rs1799929 | 8 | *NAT2* | 18400484 | Synonymous | 0.289 |
| rs1799930 | 8 | *NAT2* | 18400593 | Missense | 0.187 |
| rs1208* | 8 | *NAT2* | 18400806 | Missense | 0.305 |
| rs1799931 | 8 | *NAT2* | 18400860 | Missense | 0.128 |
| rs2294008 | 8 | *PSCA* | 142680513 | Intron | 0.322 |
| rs1927914 | 9 | *TLR4* | 117702447 | Upstream | 0.365 |
| rs8176747* | 9 | *ABO* | 133255928 | Missense | 0.046 |
| rs8176746* | 9 | *ABO* | 133255935 | Missense | 0.047 |
| rs505922 | 9 | *ABO* | 133273813 | Intron | 0.219 |
| rs10508902 | 10 | *ARHGAP22* | 48433138 | Intron | 0.324 |
| rs5030737* | 10 | *MBL2* | 52771482 | Missense | 0.025 |
| rs1057910 | 10 | *CYP2C9* | 94981296 | Missense | 0.026 |
| rs2031920 | 10 | *CYP2E1* | 133526341 | Upstream | 0.145 |
| rs1993116* | 11 | *CYP2R1* | 14888688 | Intron | 0.318 |
| rs10500804* | 11 | *CYP2R1* | 14888727 | Intron | 0.477 |
| rs12794714 | 11 | *CYP2R1* | 14892029 | Synonymous | 0.481 |
| rs2060793 | 11 | *CYP2R1* | 14893764 | 5' UTR | 0.327 |
| rs7944926* | 11 | *NADSYN1* | 71454579 | Intron | 0.45 |
| rs12800438 | 11 | *NADSYN1* | 71459957 | Intron | 0.46 |
| rs3794060* | 11 | *NADSYN1* | 71476633 | Intron | 0.448 |
| rs3829251 | 11 | *NADSYN1* | 71483513 | Intron | 0.249 |
| rs530537 | 11 | *CASP1* | 105027786 | Intron | 0.499 |
| rs4986938 | 14 | *ESR2* | 64233098 | Non Coding | 0.205 |
| rs1950902 | 14 | *MTHFD1* | 64415662 | Missense | 0.073 |
| rs2236225 | 14 | *MTHFD1* | 64442127 | Missense | 0.373 |
| rs762551 | 15 | *CYP1A2* | 74749576 | Intron | 0.236 |
| rs16260 | 16 | *CDH1* | 68737131 | Upstream | 0.266 |
| rs2297518 | 17 | *NOS2* | 27769571 | Missense | 0.137 |
| rs7220169 | 17 | *GAST* | 41715448 | Synonymous | 0.219 |
| rs713041 | 19 | *GPX4* | 1106616 | Synonymous | 0.386 |
| rs2241715* | 19 | *TGFB1* | 41350981 | Intron | 0.497 |
| rs1800469 | 19 | *B9D2* | 41354391 | Downstream | 0.5 |
| rs13042395 | 20 | *SLC52A3* | 773867 | Intron | 0.053 |
| rs6116569 | 20 | *SLC23A2* | 4884071 | Intron | 0.476 |
| rs6133175 | 20 | *SLC23A2* | 4911113 | Intron | 0.353 |
| rs6053005 | 20 | *SLC23A2* | 4977054 | Intron | 0.379 |

| | | | | | |
|---|---|---|---|---|---|
| rs17217119 | 20 | *CYP24A1* | 54126051 | none | 0.454 |
| rs225439 | 21 | *TFF3* | 42311295 | Downstream | 0.382 |

*Removed from final analysis: Not in HWE, MAF<0.5 and/or in LD

Minor Allele Frequency (MAF)
