## Supplemental Table 3 for "Ornithine Decarboxylase (*ODC1*) gene variant (rs2302615) is associated with gastric cancer independently of *Helicobacter pylori* CagA serostatus"

**Table S3. Single nucleotide polymorphisms (SNPs) Removed From Each Linkage Disequilibrium Pair**

| SNP | Chromosome | Gene | Location | Genic Location | In LD With | $R^2$ |
|---|---|---|---|---|---|---|
| rs1800872 | 1 | *IL10* | 206773062 | Upstream | rs1800871 | 0.992 |
| rs1143627 | 2 | *IL1B* | 112836810 | 5- UTR | rs16944 | 0.994 |
| rs10004195 | 4 | *TLR10* | 38783103 | Upstream | rs12233670 | 0.904 |
| rs12233670 | 4 | *TLR10* | 38785595 | none | rs4543123 | 0.943 |
| rs4833095 | 4 | *TLR1* | 38798089 | Missense | rs5743604 | 0.965 |
| rs17616434 | 4 | *TLR1* | 38811255 | none | rs5743604 | 0.944 |
| rs2282679 | 4 | *GC* | 71742666 | Intron | rs4588 | 0.984 |
| rs1155563 | 4 | *GC* | 71777771 | Intron | rs4588 | 0.938 |
| rs1800797 | 7 | *IL6* | 22726602 | Intron | rs1800795 | 0.995 |
| rs1801280 | 8 | *NAT2* | 18400344 | Missense | rs1799929 | 0.973 |
| rs1208 | 8 | *NAT2* | 18400806 | Missense | rs1799929 | 0.951 |
| rs1993116 | 11 | *CYP2R1* | 14888688 | Intron | rs2060793 | 0.974 |
| rs10500804 | 11 | *CYP2R1* | 14888727 | Intron | rs12794714 | 0.993 |
| rs7944926 | 11 | *NADSYN1* | 71454579 | Intron | rs12800438 | 0.979 |
| rs3794060 | 11 | *NADSYN1* | 71476633 | Intron | rs12800438 | 0.978 |
| rs2241715 | 19 | *TGRB1* | 41350981 | Intron | rs1800469 | -0.991 |
