## Supplemental Table 4 for "Ornithine Decarboxylase (*ODC1*) gene variant (rs2302615) is associated with gastric cancer independently of *Helicobacter pylori* CagA serostatus"

**Table S4. Unadjusted associations of single nucleotide polymorphisms (SNPs) with Gastric Cancer Risk**

| Chromosome | SNP | Gene | OR (95% CI) | p-value |
|---|---|---|---|---|
| 1 | rs1801131 | *MTHFR* | 1.03 (0.83, 1.27) | 0.824 |
| 1 | rs1801133 | *MTHFR* | 1.18 (1.01, 1.36) | 0.037 |
| 1 | rs1846522 | *FAF1* | 0.87 (0.63, 1.21) | 0.411 |
| 1 | rs1137101 | *LEPR* | 0.96 (0.82, 1.21) | 0.619 |
| 1 | rs5845 | *SELENOF* | 0.96 (0.79, 1.16) | 0.648 |
| 1 | rs1800871 | *IL10* | 1.05 (0.90, 1.22) | 0.571 |
| 1 | rs1800896 | *IL10* | 1.01 (0.84, 1.20) | 0.945 |
| 1 | rs1051740 | *EPHX1* | 0.98 (0.84, 1.15) | 0.837 |
| 2 | rs2302615 | *ODC1* | 1.21 (1.14, 1.80) | 0.027 |
| **2** | **rs1800440** | ***CYP1B1*** | **1.40 (1.08, 1.81)** | **0.011** |
| **2** | **rs1056836** | ***CYP1B1*** | **1.33 (1.12, 1.29)** | **0.001** |
| 2 | rs10197208 | *GKN3P* | 0.96 (0.82, 1.12) | 0.587 |
| 2 | rs1143634 | *IL1B* | 1.10 (0.86, 1.42) | 0.438 |
| 2 | rs16944 | *IL1B* | 0.89 (0.76, 1.03) | 0.118 |
| 2 | rs419598 | *IL1RN* | 1.02 (0.87, 1.19) | 0.811 |
| 2 | rs1063728 | *RPRM* | 1.00 (0.85, 1.18) | 0.998 |
| 2 | rs61261057 | *UGT1A10* | NA | 0.303 |
| 2 | rs17868323 | *UGT1A10* | 1.10 (0.95, 1.28) | 0.209 |
| 2 | rs11692021 | *UGT1A10* | 1.18 (0.98, 1.40) | 0.08 |
| **3** | **rs35683** | ***GHRL*** | **1.23 (1.04, 1.45)** | **0.015** |
| 3 | rs9841504 | *ZBTB20* | 0.89 (0.75, 1.05) | 0.173 |
| **4** | **rs4129009** | ***TLR10*** | **1.36 (1.07, 1.75)** | **0.014** |
| 4 | rs4543123 | *TLR1* | 1.24 (1.02, 1.49) | 0.031 |
| 4 | rs5743604 | *TLR1* | 0.92 (0.79, 1.06) | 0.25 |
| 4 | rs6815814 | *TLR1* | 1.05 (0.90, 1.22) | 0.546 |
| 4 | rs4588 | *GC* | 0.94 (0.78, 1.13) | 0.512 |
| 4 | rs7041 | *GC* | 0.98 (0.84, 1.14) | 0.788 |
| 4 | rs4073 | *CXCL8* | 1.06 (0.91, 1.24) | 0.445 |
| 5 | rs13361707 | *PRKAA1* | 1.23 (1.02, 1.49) | 0.029 |
| 5 | rs7579 | *CCDC152* | 1.02 (0.87, 1.18) | 0.846 |
| 5 | rs3877599 | *CCDC152* | 0.87 (0.87, 1.18) | 0.233 |
| 6 | rs3020314 | *ESR1* | 0.99 (0.85, 1.15) | 0.869 |
| 6 | rs4880 | *SOD2* | 1.16 (1.00, 1.36) | 0.054 |
| 7 | rs2066853 | *AHR* | 1.04 (0.84, 1.27) | 0.728 |
| 7 | rs1800795 | *IL6* | 1.03 (0.83, 1.28) | 0.778 |
| 7 | rs7799039 | *LOC105375494* | 1.07 (0.91, 1.23) | 0.424 |
| 8 | rs15561 | *NAT1* | 1.05 (0.90, 1.23) | 0.532 |
| 8 | rs1041983 | *NAT2* | 0.93 (0.80, 1.09) | 0.37 |
| 8 | rs1799929 | *NAT2* | 1.07 (0.91, 1.27) | 0.405 |
| 8 | rs1799930 | *NAT2* | 1.07 (0.88, 1.28) | 0.517 |
| 8 | rs1799931 | *NAT2* | 0.81 (0.64, 1.01) | 0.066 |
| **8** | **rs2294008** | ***PSC4*** | **1.24 (1.06, 1.46)** | **0.009** |
| **9** | **rs1927914** | ***TLR4*** | **0.75 (0.65, 0.88)** | **0.0004** |
| 9 | rs505922 | *ABO* | 0.93 (0.78, 1.11) | 0.406 |

| | | | | |
|---|---|---|---|---|
| 10 | rs10508902 | *ARHGAP22* | 0.97 (0.83, 1.14) | 0.711 |
| 10 | rs2031920 | *CYP2E1* | 0.92 (0.75, 1.14) | 0.431 |
| 11 | rs12794714 | *CYP2R1* | 1.04 (0.90, 1.21) | 0.583 |
| 11 | rs2060793 | *CYP2R1* | 1.00 (0.85, 1.17) | 0.971 |
| 11 | rs12800438 | *NADSYN1* | 0.87 (0.75, 1.01) | 0.069 |
| 11 | rs3829251 | *NADSYN1* | 1.16 (0.97, 1.38) | 0.104 |
| **11** | **rs530537** | ***CASP1*** | **0.77 (0.66, 0.90)** | **0.0007** |
| 14 | rs4986938 | *ESR2* | 1.14 (0.96, 1.39) | 0.133 |
| 14 | rs1950902 | *MTHFD1* | 1.11 (0.83, 1.50) | 0.469 |
| 14 | rs2236225 | *MTHFD1* | 0.93 (0.80, 1.08) | 0.349 |
| 15 | rs762551 | *CYP1A* | 0.91 (0.75, 1.08) | 0.268 |
| 16 | rs16260 | *CDH1* | 1.18 (0.99, 1.39) | 0.053 |
| 17 | rs2297518 | *NOS2* | 0.93 (0.75, 1.15) | 0.498 |
| 17 | rs7220169 | *GAST* | 1.05 (0.88, 1.26) | 0.584 |
| 19 | rs713041 | *GPX4* | 0.94 (0.81, 1.10) | 0.442 |
| 19 | rs1800469 | *B9D2* | 1.09 (0.78, 1.06) | 0.249 |
| 20 | rs13042395 | *SLC52A3* | 1.25 (0.89, 1.75) | 0.2 |
| 20 | rs6116569 | *SLC52A2* | 1.10 (0.95, 1.28) | 0.221 |
| 20 | rs6133175 | *SLC52A2* | 0.98 (0.84, 1.15) | 0.829 |
| 20 | rs6053005 | *SLC52A2* | 0.92 (0.78, 1.08) | 0.321 |
| 20 | rs17217119 | *CYP24A1* | 1.05 (0.90, 1.23) | 0.54 |
| 21 | rs225439 | *TFF3* | 0.91 (0.78, 1.07) | 0.255 |

Bolded SNPs passed False Discovery Rate (FDR) cut off
