## Supplemental Table 5 for "Ornithine Decarboxylase (*ODC1*) gene variant (rs2302615) is associated with gastric cancer independently of *Helicobacter pylori* CagA serostatus"

**Table S6. Adjusted associations of single nucleotide polymorphisms (SNPs)**

| Chromosome | SNP | Gene | OR* | p-value |
|---|---|---|---|---|
| 1 | rs1801131 | *MTHFR* | | 0.885 |
| **1** | **rs1801133** | ***MTHFR*** | **1.18 (0.99, 1.39)** | **0.058** |
| 1 | rs1846522 | *FAF1* | | 0.226 |
| 1 | rs1137101 | *LEPR* | | 0.691 |
| 1 | rs5845 | *SELENOF* | | 0.61 |
| 1 | rs1800871 | *IL10* | | 0.8 |
| 1 | rs1800896 | *IL10* | | 0.837 |
| 2 | rs2302615 | *ODC1* | N/A | N/A |
| **2** | **rs1800440** | ***CYPTB1*** | **1.22 (1.03, 1.46)** | **0.047** |
| **2** | **rs1056836** | ***CYPTB1*** | **1.22 (1.03, 1.45)** | **0.0473** |
| 2 | rs10197208 | *GKN3P* | | 0.79 |
| 2 | rs1143634 | *IL1B* | | 0.237 |
| 2 | rs1143627 | *IL1B* | | 0.43 |
| 2 | rs16944 | *IL1B* | | 0.377 |
| 2 | rs419598 | *IL1RN* | | 0.854 |
| 2 | rs1063728 | *RPRM* | | 0.796 |
| 2 | rs61261057 | *UGT1A10* | | 0.971 |
| 2 | rs17868323 | *UGT1A10* | | 0.365 |
| 2 | rs11692021 | *UGT1A10* | | 0.45 |
| **3** | **rs35683** | ***GHRL*** | **0.82 (0.67, 1.00)** | **0.079** |
| 3 | rs9841504 | *ZBTB20* | | 0.154 |
| **4** | **rs4129009** | ***TLR10*** | **0.78 (0.59, 1.03)** | **0.079** |
| 4 | rs4543123 | *TLR1* | | 0.319 |
| 4 | rs5743604 | *TLR1* | | 0.443 |
| 4 | rs6815814 | *TLR1* | | 0.8 |
| 4 | rs4588 | *GC* | | 0.789 |
| 4 | rs7041 | *GC* | | 0.799 |
| 4 | rs4073 | *CXCL8* | | 0.94 |
| **5** | **rs13361707** | ***PRKAA1*** | **1.24 (1.00, 1.53)** | **0.048** |
| 5 | rs7579 | *CCDC152* | | 0.73 |
| 5 | rs3877899 | *CCDC152* | | 0.363 |
| 6 | rs3020314 | *ESR1* | | 0.624 |
| 6 | rs4880 | *SOD2* | | 0.173 |
| 7 | rs2066853 | *AHR* | | 0.56 |
| 7 | rs1800795 | *IL6* | | 0.509 |
| 8 | rs5030839 | *NAT1* | | 0.302 |
| 8 | rs15561 | *NAT1* | | 0.789 |
| 8 | rs1041983 | *NAT2* | | 0.113 |
| 8 | rs1799929 | *NAT2* | | 0.252 |
| 8 | rs1799930 | *NAT2* | | 0.599 |
| 8 | rs1799931 | *CYP2C9* | | 0.198 |
| 8 | rs2294008 | *PSCA* | | 0.168 |

| | | | | |
|---|---|---|---|---|
| **9** | **rs1927914** | ***TLR4*** | **1.22 (1.03, 1.46)** | **0.023** |
| 9 | rs505922 | *ABO* | | 0.332 |
| 10 | rs10508902 | *ARHGAP22* | | 0.944 |
| 10 | rs2031920 | *CYP2E1* | | 0.331 |
| 11 | rs12794714 | *CYPR1* | | 0.34 |
| 11 | rs2060793 | *CYPR1* | | 0.575 |
| 11 | rs12800438 | *NADSYN1* | | 0.264 |
| 11 | rs3829251 | *NADSYN1* | | 0.15 |
| **11** | **rs530537** | ***CASP1*** | **1.22 (1.03, 1.45)** | **0.02** |
| **14** | **rs4986938** | ***ESR2*** | **1.22 (0.99, 1.51)** | **0.069** |
| 14 | rs1950902 | *MTHFD1* | | 0.627 |
| 14 | rs2236225 | *MTHFD1* | | 0.483 |
| 15 | rs762551 | *CYP1A2* | | 0.597 |
| 16 | rs16260 | *CDH1* | | 0.238 |
| 17 | rs2297518 | *NOS2* | | 0.911 |
| 17 | rs7220169 | *GAST* | | 0.951 |
| 19 | rs713041 | *GPX4* | | 0.876 |
| 19 | rs1800469 | *B9D2* | | 0.88 |
| **20** | **rs13042395** | ***SLC52A3*** | **1.39 (0.96, 2.03)** | **0.087** |
| 20 | rs6116569 | *SLC23A2* | | 0.388 |
| 20 | rs6133175 | *SLC23A2* | | 0.668 |
| 20 | rs6053005 | *SLC23A2* | | 0.416 |
| 20 | rs17217119 | *CYP24A1* | | 0.523 |
| 21 | rs225439 | *TFF3* | | 0.172 |

*Odds ratio only included if p <0.10 (shown in bold), adjusted for age, sex, CagA serostatus, and ODC genotypes
