## Supplemental Figure 1 for "Ornithine Decarboxylase (*ODC1*) gene variant (rs2302615) is associated with gastric cancer independently of *Helicobacter pylori* CagA serostatus"

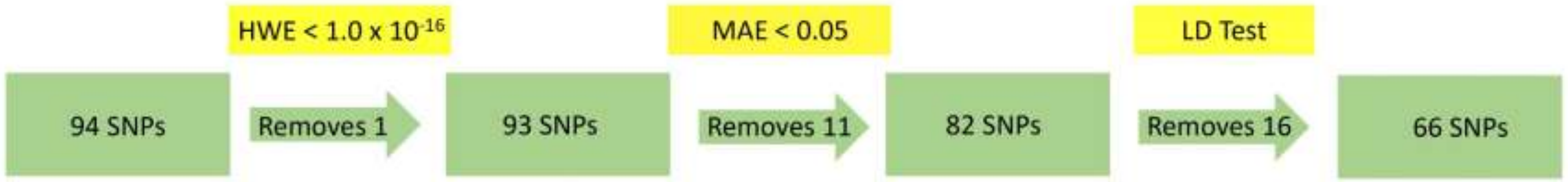


**Supplemental Figure 1:** The original 94 SNPs were pruned using Hardy-Weinberg equilibrium ($p < 1.0 \times 10^{-16}$) which removed SNP rs4072037 due to potential genotyping error. In addition, a minor allele frequency test (< 0.05) removed 11 SNPs and a Linkage Disequilibrium (LD) detected 20 SNP pairs in which 4 SNPs appeared in two pairs, removing 16 SNPs. SNPs were removed due to LD with a threshold of R^2 >= 0.9. Therefore, 66 SNPs remained in the final analysis.
